# Multi-Omics Integration of Transcriptomics and Metabolomics with Machine Learning Uncovers Novel Risk Factors for Alzheimer’s disease

**DOI:** 10.64898/2026.02.28.26347204

**Authors:** Jerome J. Choi, Corinne D. Engelman, Tianyuan Lu

**Affiliations:** Department of Population Health Sciences, University of Wisconsin-Madison; Department of Biostatistics and Medical Informatics, University of Wisconsin-Madison; Center for Genomic Science Innovation; Center for Demography of Health and Aging; Center for Precision Medicine

**Keywords:** Alzheimer’s disease, machine learning, multi-omics, biomarkers

## Abstract

**Background:** Alzheimer’s disease (AD) is a neurodegenerative disorder marked by cognitive decline, memory impairment, and functional deterioration. Its complex pathogenesis involves factors such as amyloid plaques, tau tangles, neuroinflammation, and synaptic dysfunction, but the precise mechanisms remain unclear, hindering effective treatment. Genetic, environmental, and lifestyle factors contribute to AD risk, yet their interactions are poorly understood. Recent advances in transcriptomics and metabolomics have shed light on the molecular underpinnings of AD, with gene expression alterations and metabolic disruptions implicated in disease progression. These multi-omics disruptions highlight the need for integrative analytical approaches to better characterize AD-relevant biology and advance biomarker discovery.

**Objectives:** To integrate genetically imputed whole blood transcriptomics and plasma metabolomics to predict cognitive performance (PACC3) and to identify risk genes and metabolites contributing to prediction, thereby characterizing molecular signatures associated with cognitive performance in AD.

**Methods:** This study applies a machine learning algorithm to integrate genetically imputed whole blood transcriptomics and measured plasma metabolomics data to predict cognitive performance, as measured by PACC3 score, using data from the Wisconsin Registry for Alzheimer’s Prevention (WRAP) cohort (N = 1,046). After training a machine learning model on WRAP, the predictive performance was evaluated using an independent dataset from the Wisconsin Alzheimer’s Disease Research Center (ADRC) cohort (N = 85). Feature importance was assessed to identify genes and metabolites that may play a role as potential risk factors in AD.

**Results:** The machine learning model achieved a normalized root mean squared error (NRMSE) of 0.743 ± 0.037 and an R² of 0.311 ± 0.016 across 5-fold holdout test folds in WRAP (p = 5.93 × 10^−30^), and an NRMSE of 0.915 and an R² of 0.061 when applied to the Wisconsin ADRC cohort. Feature importance revealed transcriptomic biomarkers such as *RIPK1*, *IL6ST*, and *BIN1* whose higher imputed expression levels were associated with poorer cognitive performance whereas other potential biomarkers including *UGP2*, *NDUFB5*, and *TMOD2* were associated with better cognitive performance, reflecting mitochondrial energy metabolism and molecular processes associated with cognitive resilience. Several predictive metabolites including benzoate, 3-phenylpropionate, and imidazolelactate also mapped to AD vulnerability signatures, while acyl-carnitine species such as hexanoylcarnitine (C6) and propionate-related metabolites aligned with metabolic resilience.

**Conclusion:** Integrated analysis of transcriptomics and metabolomics demonstrated potential utility for identifying candidate biomarkers associated with cognition in AD. Genes and metabolites reflecting inflammatory signaling, mitochondrial dysregulation, and lipid metabolism emerged consistently among the most influential contributors. These findings align with well-established AD vulnerability pathways and highlight convergent biology across two omics layers. Collectively, this supports the value of multi-omics integration for improving molecular characterization of AD and advancing biomarker prioritization for future mechanistic and translational studies.

## Introduction

Alzheimer’s disease (AD) is a neurodegenerative disease characterized by progressive cognitive decline, memory impairment, and functional deterioration, representing a major public health challenge(1). The pathogenesis of AD is multifactorial, involving the accumulation of amyloid plaques, neurofibrillary tangles, neuroinflammation, and synaptic dysfunction.(2–5) Despite significant advances in understanding the molecular underpinnings of the disease, the precise mechanisms remain unclear, and the complexity of AD presents a challenge for early diagnosis and effective therapeutic development. Even though numerous genetic, environmental, and lifestyle factors have been associated with AD risk,(6,7) the specific biomarkers underlying this risk remain unclear. As a result, there is a pressing need for more comprehensive approaches to elucidate the molecular landscape of AD and identify novel biomarkers that could aid in early detection and intervention, given that AD pathology can begin decades before clinical symptoms emerge.(8)

In recent years, transcriptomics and metabolomics have emerged as key omics approaches for investigating the molecular basis of AD.(9) Transcriptomics, which analyzes gene expression patterns, provides insights into the dysregulated cellular processes in AD, such as neuronal loss, synaptic dysfunction, and inflammatory responses.(10) Alterations in the expression of genes related to amyloid processing, tau metabolism, and neuroinflammation have been linked to disease progression, highlighting the role of transcriptional dysregulation in AD pathophysiology.(11) When tissue-specific RNA sequencing data are unavailable, genetically imputed transcriptomics provides a cost-effective and scalable alternative by leveraging genotype–expression reference panels to infer gene expression in cohorts with genetic data. Methods such as PrediXcan(12) enable systematic investigation of genetically regulated gene expression and its association with disease risk in population-scale studies. On the other hand, metabolomics focuses on the small molecules involved in cellular metabolism, offering a unique window into the biochemical changes that occur in AD.(13)

Studies have shown that metabolites associated with energy metabolism, oxidative stress, and neurotransmitter pathways are altered in AD, suggesting that disrupted metabolic networks contribute to disease onset and progression.(14) While both transcriptomics and metabolomics provide valuable insights into AD, the interplay between genes and metabolites suggests that a more integrative approach is required.(15)

Multi-view integration of omics data provides a comprehensive understanding of systems biology, particularly in complex diseases such as AD.(16,17) By combining data from multiple omics layers, it is possible to capture a more holistic view of the molecular mechanisms underlying AD. Moreover, this integrative approach may allow for the identification of key biomarkers and pathways that may be missed when examining a single omics layer in isolation.(18) This study applies a machine learning method, Cooperative learning(19) to integrate genetically imputed transcriptomic data and measured metabolomic data, predict cognitive performance as measured by the Preclinical Alzheimer’s Cognitive Composite 3 (PACC3) score, and identify potential AD-risk factors from the Wisconsin Registry for Alzheimer’s Prevention (WRAP)(20) cohort. Data from the Wisconsin Alzheimer’s Disease Research Center (ADRC) are used for independent validation.

## Methods

### Study design

An overview of the study design is presented in **FIGURE 1**. Gene expression data were imputed using the genotypes of WRAP and Wisconsin ADRC participants via PrediXcan(12). To reduce the dimensionality, untargeted transcriptomic and metabolomic analyses were performed, including age, sex, *APOE* score, and years of education as covariates, to predict PACC3 in WRAP using linear regression. Subsequently, nominally significant genes and metabolites (p-value < 0.05) were input into a machine learning (ML) model, which was trained to predict the outcome by implementing Cooperative learning. After training the ML model, biomarker importance for individual genes and metabolites were computed. Lastly, the Wisconsin ADRC cohort was used to independently validate the prediction performance of the ML model.

**FIGURE 1.**
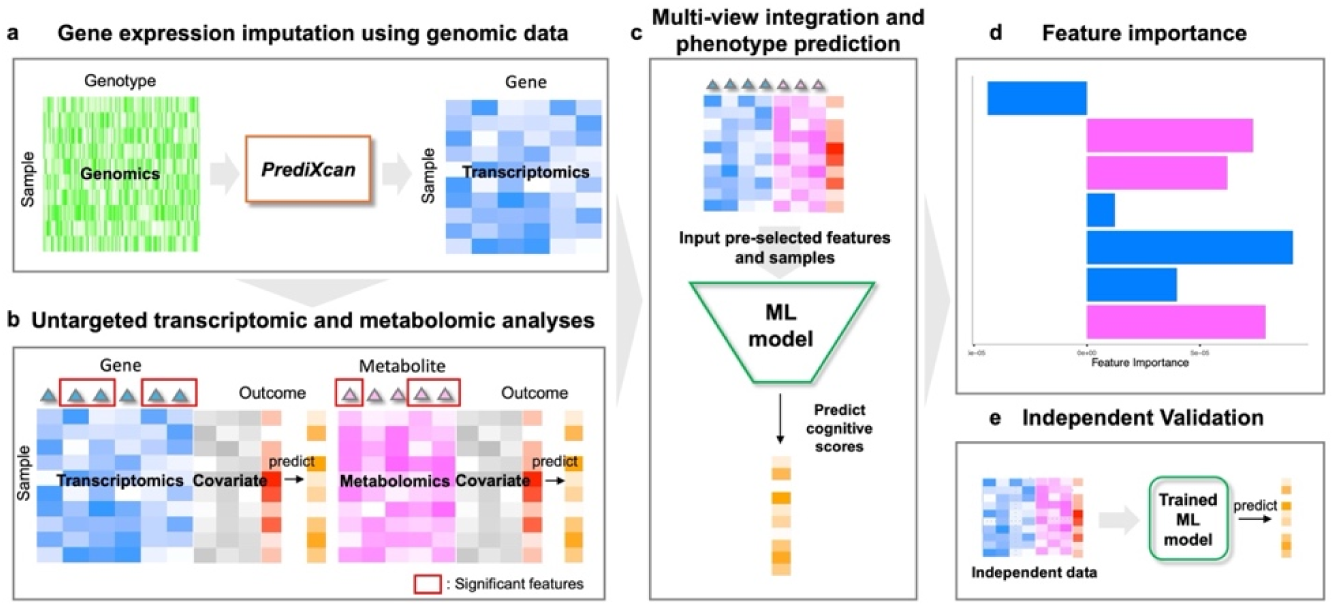
Workflow diagram. **a** Gene expression data for the WRAP participants is imputed using PrediXcan. **b** Untargeted transcriptomics and metabolomics adjusting for covariates are implemented to pre-select significant biomarkers. **c** Multi-omics data are integrated and cognitive scores are predicted using a machine learning (ML) model. **d** Biomarker importance values are computed by a ML model. **e** Independent validation for predicting AD phenotypes using the Wisconsin ADRC cohort as independent multi-view data is performed.

### Study participants and cohort description

The cohort for the main analysis, WRAP, is an ongoing longitudinal study designed to identify midlife factors associated with the development of AD. Participant enrollment began in 2001, with the first follow-up visit occurring 2 to 4 years after the baseline visit, and all subsequent visits taking place at 2-year intervals thereafter. WRAP participants were dementia-free at enrollment (mean age 54 years). All study procedures were approved by the University of Wisconsin School of Medicine and Public Health Institutional Review Board and are in accordance with the Declaration of Helsinki.

Wisconsin ADRC participants were recruited from multiple sources such as memory clinic providers, events, media, partner organizations, and other studies. Adults aged 45 years or older with decisional capacity and English fluency were eligible for enrollment. Exclusion criteria included active major medical or psychiatric illnesses and lack of a study partner. This study included participants who have normal cognitive status based on their diagnoses.

### Alzheimer’s disease outcomes

PACC3 score is a global cognitive composite score used to assess the cognitive performance of participants in the WRAP study. During each study visit, participants complete a detailed cognitive battery, which is described in more detail elsewhere(20). Longitudinal global cognitive performance is evaluated using a three-test version of the modified PACC(21), which includes the Rey Auditory Verbal Learning Test (AVLT; Trials 1–5)(22), the Wechsler Memory Scale Logical Memory II(23), and the Wechsler Adult Intelligence Scale-Revised Digit Symbol Substitution.(24) The Logical Memory II test was introduced at Visit 2 for most participants (94%), so the PACC3 baseline is generally at Visit 2, with Visit 1 being the baseline for recent enrollees where it was administered at Visit 1. Each test score is converted into a Z-score, using the mean and standard deviation from baseline scores of cognitively unimpaired individuals.

The Wisconsin ADRC uses a slightly different set of neuropsychological tests compared to WRAP. To maximize the use of available data, we collaborated with neuropsychologists at the Wisconsin ADRC to create a PACC-3-Trail Making Test (TMT) score for the replication analysis. Specifically, we converted the Craft Story score into an estimated Logical Memory score using a published crosswalk table and followed previous methodology by substituting the Digit Symbol score with the total completion time from the TMT-B test.(21,25). Since the crosswalk table only provides a conversion for the Craft Story score to the Logical Memory score for the first five visits, we limited the replication analysis to data from these five visits. The final PACC-3-TMT score was calculated by standardizing and averaging the results from the AVLT, the estimated Logical Memory score, and the reversed TMT-B test scores.

### Plasma metabolite collections and quality control

#### Collection

Participants underwent venipuncture, most after fasting ≥8 hours, and provided 30 mL of blood into 3×10 mL lavender top EDTA tubes (BD 366643; Franklin Lakes, New Jersey, USA). Samples were mixed gently by inverting 10–12 times and were centrifuged 15 min at 2000 g at room temperature within 1 h of collection. Plasma samples were aliquoted into 2 mL cryovials (Wheaton Cryoelite W985863; Millville, New Jersey, USA). Aliquoted plasma was frozen at −80°C within 90 min and stored until overnight shipment to Metabolon, Inc. (Durham, NC), which similarly kept samples frozen at −80° C until analysis. Participants who fasted <8 hours prior to venipuncture were excluded (n = 298).

#### Quality control

Samples with missing values for more than 40% of metabolites were removed (0 plasma samples) prior to analysis. Non-xenobiotic metabolites with missing values for >75% (n=43), and xenobiotic metabolites with missing values for >99% (n=75) of all samples were removed.

#### Fasting status

Samples from individuals who fasted less than 8 hours prior to venipuncture, or who did not provide fasting status were removed.

### Genotyping, quality control, and imputation

#### Genotyping array data

DNA was extracted from whole blood samples and genotyped at the University of Wisconsin Biotech Center in three batches (2017, 2021, 2024) using the Illumina Multi-Ethnic Genotyping Array (MEGA).

#### Processing and Quality Control

Initial quality control was performed separately for each batch using PLINK v1.9(26). SNPs that did not map to any chromosomes were removed, followed by variants missing in >5% of samples and samples with >5% missing variants. Additional filters were applied to remove variants missing in >2% of remaining samples and samples with >2% missing variants. SNPs with canonical nucleotide alleles (A, C, T, or G) were retained, and sex discrepancies were checked using X chromosome homozygosity, removing 10 samples with inconsistent reported sex. The data were remapped from hg37 to hg38 using the UCSC liftover tool(27). The HRC checker tool was used to process the data before imputation, removing duplicates, mismatches, and monomorphic SNPs. Data were sorted by chromosome and saved as VCF files using BCFtools(28). Imputation was done via the TOPMed Imputation Server using Eagle v2.4 and the TOPMed reference panel r3(29–31). After imputation, variants with a low INFO score (R^2^ <0.8) were removed. Data were merged and filtered for genotyping rate >98% and MAF >0.001. The final dataset consisted of 14,980,298 SNPs.

#### Relatedness and genetic ancestry

A pruned set of the genotyped SNPs, using PLINK v1.9 --indep-pairwise 50 10 0.1, was used to estimate relatedness and genetic similarities to 1000 Genomes superpopulations(32). Kinship estimation was performed using KING v2.3.1(33) to identify genetically identical individuals and infer first-degree familial relationships, which were then used to define subsets of related and unrelated samples for downstream ancestry and population structure analyses. The pruned SNPs, in a subset of unrelated samples, were used in a supervised learning model in ADMIXTURE v1.3.0, using the default maximum likelihood estimation, to determine proportions of genetic similarities between the Wisconsin samples and the 1000 Genomes defined continental populations (African, Admixed American, East Asian, European, and South Asian). The subset of related samples was projected onto the resulting population structure output from ADMIXTURE(34).

#### *APOE* score

The *APOE* score is a weighted risk score for the effect of the six *APOE* genotypes on AD neuropathology. It is calculated using the natural log (ln) of the odds ratios (OR) for the six different *APOE* genotypes, based on a study of AD neuropathology(35). The *APOE* genotypes considered are ε2ε2, ε2ε3, ε3ε3, ε2ε4, ε3ε4, and ε4ε4, with ε3ε3 serving as the reference group.(36)

#### Gene expression imputation

The recently published pre-trained database, which used GTEx whole blood data, was obtained from the PredictDB Data Repository(37). Genotypes from WRAP and Wisconsin ADRC participants were used to impute whole blood gene expression levels for the participants via PrediXcan. Specifically, genetic risk scores for predicting tissue-specific gene expression levels were pre-trained based on GTEx, which contains measured individual-level genetics and transcriptomics data. PrediXcan then calculates genetically predicted whole blood gene expression levels using genotypes of WRAP and Wisconsin ADRC participants, although transcriptomics data for these participants were not measured.

### Statistical analyses

#### Sample selection

WRAP participants were selected based on the availability of plasma metabolite measurements, genotypes, outcome data, and covariates such as age, sex, years of education, and *APOE* score. Among multiple visits, we selected the last observation for each participant, where the outcome was measured at or within six months after the plasma metabolites. To reduce confounding due to population stratification, we restricted the analyses to participants with likely European genetic ancestry (>0.5 admixture). For the Wisconsin ADRC cohort, participants diagnosed with dementia were excluded.

We randomly assigned the WRAP participants into a training dataset (75%) and a holdout test dataset (25%).

#### Pre-selecting potential AD-risk factors

We fitted a linear regression model for each biomarker (6,678 imputed gene expression features and 1,125 plasma metabolites) and the PACC3 outcome in the WRAP training set. For each biomarker, the model included the biomarker of interest together with the predefined covariates, to account for demographic, genetic, and educational factors known to influence cognitive performance:

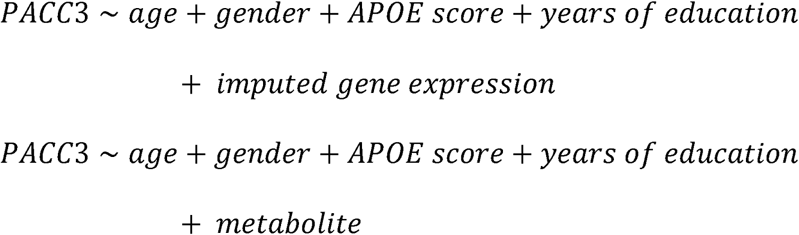

Imputed gene expression levels and metabolites with a p-value < 0.05 were considered nominally significant biomarkers. These untargeted imputed transcriptomic and metabolomic analyses were performed to improve the computational efficiency of downstream machine learning modelling by focusing on potentially relevant associations.

#### Predictive modeling

##### Baseline Model

A baseline prediction model for cognitive performance was constructed in the WRAP training set, including age at assessment, sex, APOE score, and years of education as predictors:

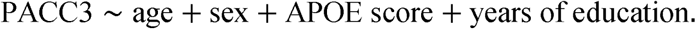

The model was trained using ordinary least squares on the training set and then applied to an independent test set to generate out-of-sample predictions of PACC3 scores. Model performance was evaluated in the test set using NRMSE. The NRMSE standardizes the error by normalizing it with respect to the interquartile range (IQR) of the observed data. The equation for NRMSE using IQR is given by:

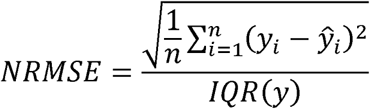

Where *y_i_* represents the actual values, ŷ*_i_* denotes the predicted values, and n is the number of participants. The term *IQN*(*y*) refers to the interquartile range of actual values, which is calculated as *Q*_3_ - *Q_l_*, where *Q*_3_ and *Q*_l_ are the third and first quartiles of the actual values, respectively.

This baseline model provides a reference for predictive performance using covariates alone, against which models incorporating omics-derived predictors were compared.

##### Machine Learning Model

Significant biomarkers, pre-selected from untargeted imputed transcriptomics and metabolomics in WRAP, were input into the multi-omics models using Cooperating Learning. The Cooperative Learning framework(19) jointly estimates multi-view regression models with view-specific regularization and a cooperative penalty that encourages shared predictive structure across omics modalities.

The model was trained using a 5-fold cross-validation, where each fold was used as a hold-out validation split to select the optimal cooperation parameter (rho) and regularization parameter (lambda). Data standardization was performed within each training fold only, ensuring no information leakage from validation or holdout samples. For each fold, the model was fit across the full lambda path, and the combination of rho and lambda that minimized validation mean squared error (MSE) was selected. The selected fold-specific model was then evaluated on the same WRAP holdout test set, resulting in five independent holdout performance estimates.

Feature importance was derived from the non-zero coefficients extracted at the selected lambda for each fold, allowing identification of influential biomarkers separately within each omics view. Following cross-validation on WRAP, the single fold that achieved the lowest holdout MSE was used as the deployed model and applied to an independent ADRC cohort for external validation to evaluate generalizability. The prediction performance of the models was assessed using NRMSE.

To assess whether model predictions were statistically associated with observed cognitive outcomes, we evaluated the linear association between predicted and observed PACC3 scores in the evaluation set using Pearson correlation. Statistical significance was assessed using a two-sided Pearson correlation test, and the corresponding p-value was reported.

#### Feature importance and enrichment analysis

Individual feature importance within each omics data type in WRAP was based on model coefficients obtained from the training set. The genes and metabolites from the best model with non-zero coefficients were considered for enrichment analyses.

For enrichment analyses, we selected the top 50 genes and top 50 metabolites with the most positive feature importance values and the top 50 with the most negative values. Gene set enrichment was performed using clusterProfiler(38) and metabolite enrichment was performed using MetaboAnalystR(39).

## Results

### Descriptive statistics for the participants

Participants from the WRAP cohort (n = 1,046) and the Wisconsin ADRC cohort (n = 85) were included in the analysis (TABLE 1). In WRAP, the mean age at assessment was 67.04 years (SD = 6.89), whereas ADRC participants were slightly older on average, with a mean age of 69.14 years (SD = 7.65). The WRAP cohort had a higher proportion of female participants, with 716 females (68.5%) and 330 males (31.5%), compared with the ADRC cohort, which included 50 females (58.8%) and 35 males (41.2%).

**TABLE 1.** Demographic of participants in two cohorts.

| Cohort | WRAP | Wisconsin<br>ADRC |
| --- | --- | --- |
| Total participants | 1,046 | 85 |
| Age (mean [SD]) | 67.04 (6.89) | 69.14 (7.65) |
| Females | 716<br>(68.5%) | 50<br>(58.8%) |
| Males | 330<br>(31.5%) | 35<br>(41.2%) |
| *APOE* score (mean [SD]) | 0.63 (1.04) | 0.61 (1.03) |
| Years of education<br>(mean [SD]) | 15.80 (2.23) | - |
| PACC3 (mean [SD]) | 0.016 (0.83) | 0.425 (0.812) |

Mean APOE scores were similar across cohorts, with an average of 0.63 (SD = 1.04) in WRAP and 0.61 (SD = 1.03) in ADRC, indicating comparable distributions of APOE scores. Years of education were available for WRAP participants only, with a mean of 15.80 years (SD = 2.23), and were not available for ADRC participants.

With respect to cognitive performance, mean PACC3 scores differed between cohorts. WRAP participants had a mean PACC3 score of 0.016 (SD = 0.83), whereas ADRC participants had a higher mean PACC3 score of 0.425 (SD = 0.812). These differences may reflect variation in cohort composition and assessment context rather than direct comparability of cognitive performance across cohorts.

### Untargeted imputed transcriptomics and metabolomics

Out of 6,678 genes with imputed whole blood expression levels, 354 were retained for downstream machine learning modeling (nominal p-value < 0.05). Meanwhile, among 1,125 measured plasma metabolites, 150 were retained (**FIGURE 2**).

**FIGURE 2.**
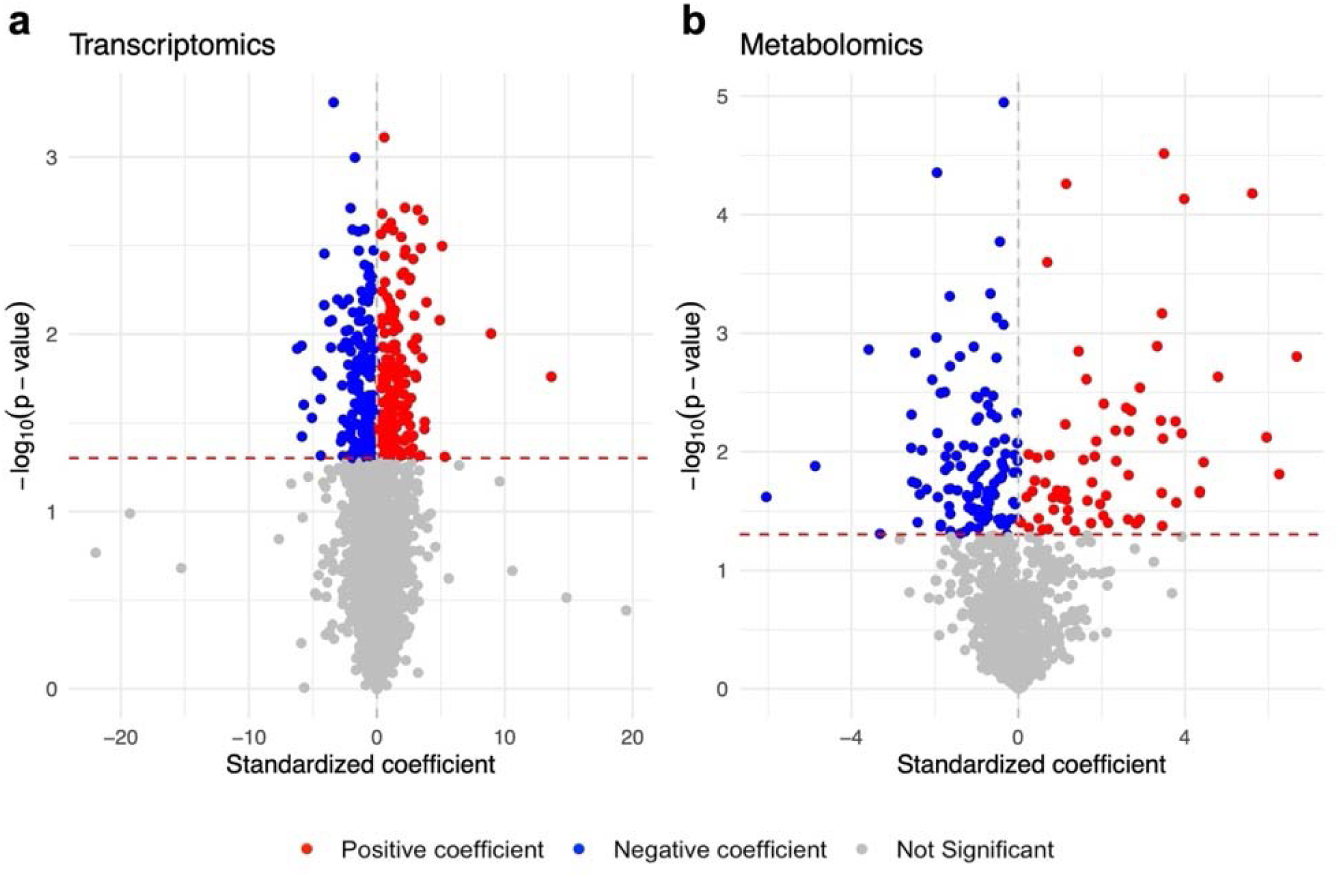
Volcano plots summarizing results of untargeted imputed transcriptomic and metabolomic analyses. P-values (y-axis) and magnitudes of associations (x-axis) for **a** untargeted imputed transcriptomics predicting PACC3, **b** untargeted metabolomics predicting PACC3. The red dashed horizontal line indicates the statistical significance threshold (−logl_l_(p) corresponding to p = 0.05), with features above the line considered statistically significant.

### AD phenotype prediction

For the main analysis, models were trained and tuned using 5-fold cross-validation on the training set (75% of the WRAP cohort), and final model performance was evaluated on an independent holdout test set comprising the remaining 25% of participants. Based on cross-validated predictions within the WRAP training set, the machine-learning model achieved a mean NRMSE of 0.743 ± 0.037 and a mean R^2^ of 0.311 ± 0.016 (TABLE 2). The predicted PACC3 values were significantly correlated with observed PACC3 values (Pearson correlation p-value = 9.911601e-25).

**TABLE 2.**
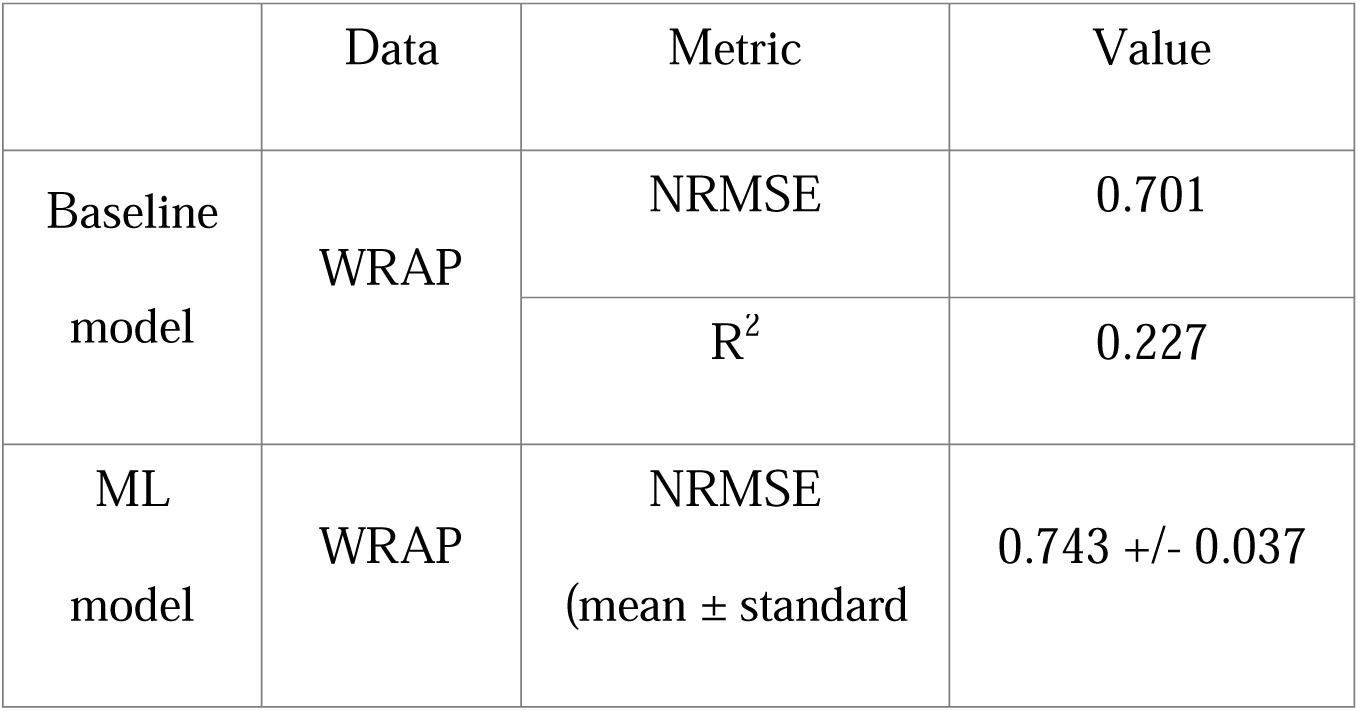
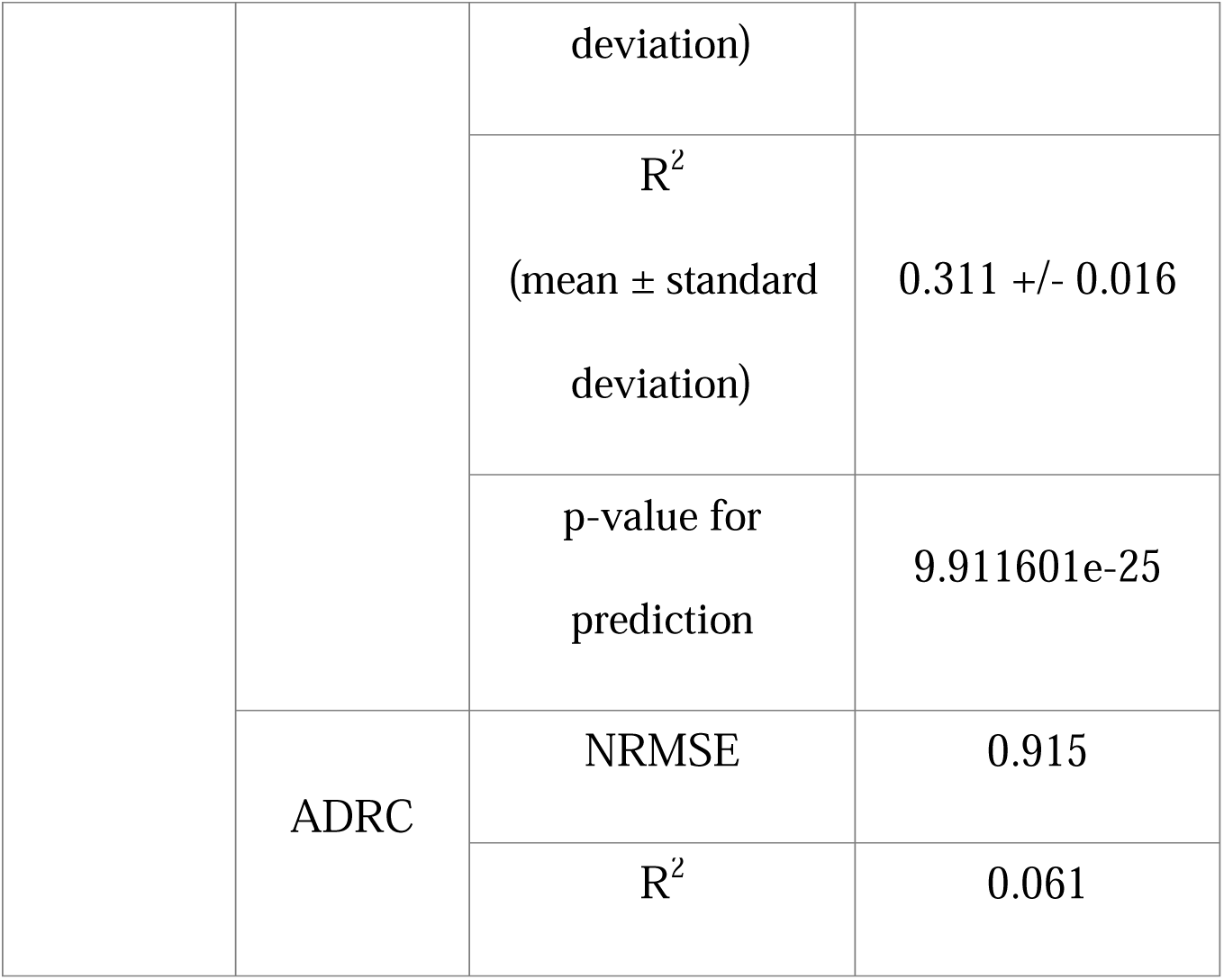
Model performance metrics across WRAP and ADRC cohorts.

As an external validation, we apply the trained model to the Wisconsin ADRC cohort, which yielded an NRMSE of 0.915 and an R² of 0.061 (TABLE 2).

### Biomarker importance

The top 30 most important transcriptomic biomarkers and metabolomic biomarkers, defined based on model coefficients, are illustrated in **Supplementary FIGURE 1a** and **Supplementary FIGURE 1b**, respectively. Biomarkers with positive coefficients indicate that higher expression is associated with better cognitive performance, whereas biomarkers with negative coefficients indicate that higher expression is associated with lower cognitive performance, consistent with AD-related vulnerability.

Genes with positive feature importance are enriched for GO biological processes related to chromatin regulation, mitochondrial proton handling, ketone utilization, and homocysteine (**FIGURE 3a**). These pathways reflect preserved neuronal energy efficiency(40), methylation capacity(41), and metabolic flexibility(42), all of which have been implicated as protective mechanisms in cognitive resilience and reduced AD progression risk.

**FIGURE 3.**
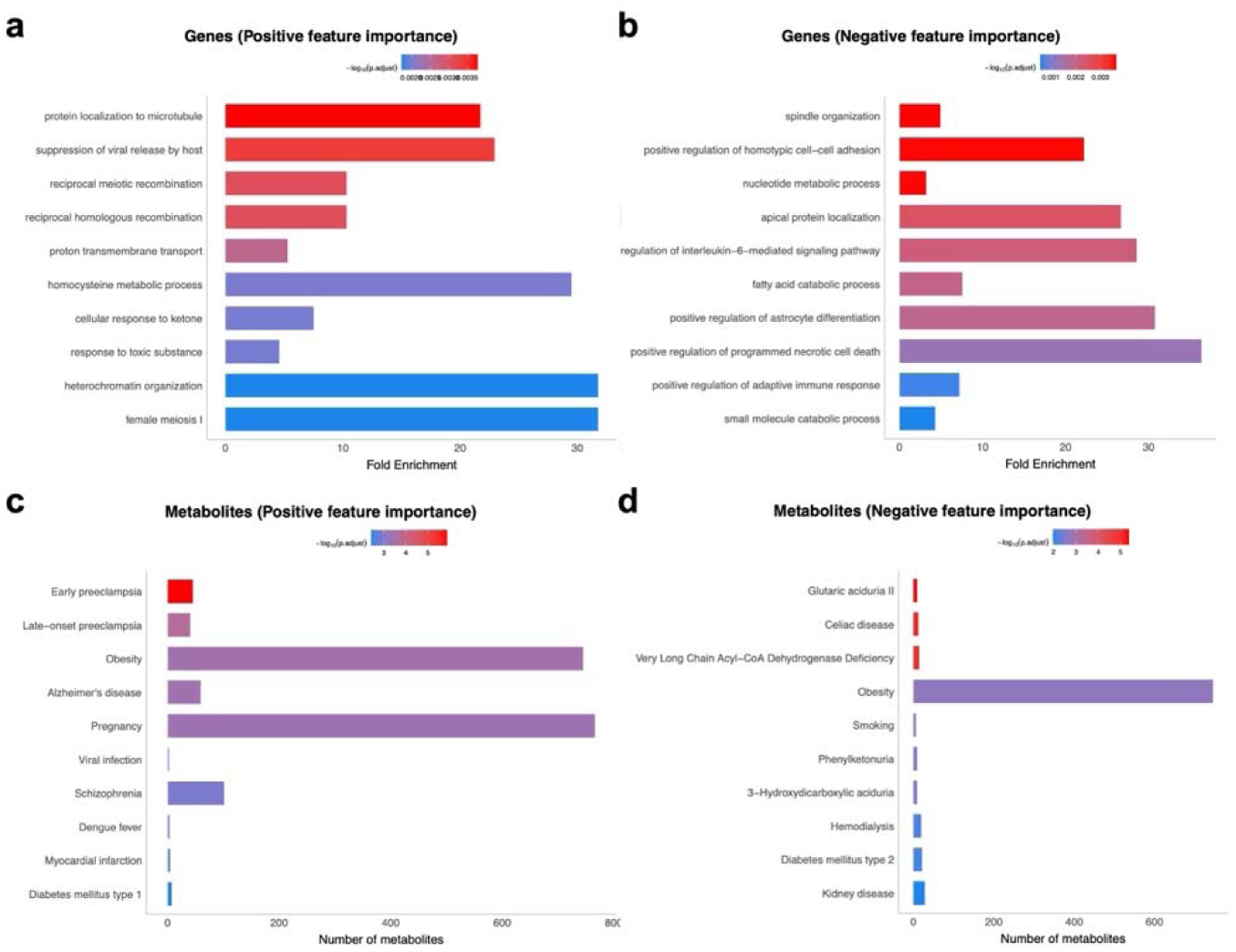
Enrichment analyses for genes and metabolites. (a) GO biological process enrichment for genes with positive feature importance. (b) GO biological process enrichment for genes with negative feature importance. In panels (a) and (b), fold enrichment indicates the over-representation of genes annotated to a given GO term relative to background expectation, with color denoting adjusted p-values. (c) Disease enrichment for metabolites with positive feature importance. (d) Disease enrichment for metabolites with negative feature importance. In panels (c) and (d), bar length represents the number of metabolites contributing to each disease category, with color indicating enrichment significance.

By contrast, genes with negative feature importance highlight inflammatory cytokine signaling, necroptotic cell death programs, and astroglial activation (**FIGURE 3b**). These immune-driven and glial-activation pathways are well-established key drivers in AD pathobiology(43), aligning with pro-inflammatory states(44), microglial/astrocyte reactivity(45), and neuronal death processes(46) that accelerate cognitive decline.

Metabolites with positive feature importance overlap with disease-associated metabolic clusters previously observed in large-scale AD and obesity metabolomics datasets (**FIGURE 3c**). These enrichments indicate that systemic metabolic traits(47), when shifted toward more efficient substrate use or balanced metabolic buffering, may promote more favorable cognitive trajectories in aging.

In contrast, metabolites with negative feature importance converge on signatures of cellular energy impairment(48), defective mitochondrial fatty-acid oxidation(49), and metabolic disease phenotypes such as diabetes and inborn errors of metabolism(50) (**FIGURE 3d**). These disrupted metabolic axes align closely with AD vulnerability biology, as impaired mitochondrial function and abnormal lipid handling are hallmarks of accelerated neurodegenerative decline(49).

## Discussion

Given the complex, multifactorial nature of AD, integrating multi-omic data offers a powerful approach to enable the identification of molecular signatures that may underlie disease susceptibility, progression, and resilience. In this study, we aimed to predict a critical AD-related phenotype, PACC3 score using imputed transcriptomic data and measured metabolomic data, via a machine learning approach, in the WRAP cohort. We then assessed the individual biomarker importance. To validate the prediction performance of the trained models, we conducted independent validation by applying the same models to predict outcomes in the Wisconsin ADRC cohort. These analyses provide valuable insights into potential AD risk factors and highlights key molecular features.

As expected, predictive performance was lower in the Wisconsin ADRC compared with WRAP, likely reflecting differences in cohort characteristics such as cognitive status and sample collection protocols. Importantly, despite this attenuation, the model retained reasonable predictive ability, suggesting that the key molecular features identified in WRAP capture underlying biological processes relevant to cognitive performance across populations. This partial generalizability supports the robustness of our multi-omic approach and highlights the potential for identifying cross-cohort biomarkers.

Enrichment analysis of genes with positive feature importance highlights pathways involved in chromatin organization, mitochondrial proton transport, ketone metabolism, and homocysteine processing. Genes such as *HP1BP3*(51) and *MTHFR*(52) implicate chromatin remodeling and one-carbon metabolism, processes essential for maintaining epigenetic regulation and methylation capacity, which have been linked to cognitive resilience. Meanwhile, mitochondrial genes including *NDUFB5*(53), *NDUFA9*(53), and *NNT*(54) converge on proton transmembrane transport and mitochondrial efficiency, suggesting that sustained bioenergetic capacity supports better cognitive outcomes. Collectively, these positive-direction genes point toward a coordinated network that maintains cellular energy homeostasis and epigenetic stability, highlighting potential mechanisms through which certain molecular profiles may buffer against age-related cognitive decline. These findings suggest that interventions supporting mitochondrial efficiency or one-carbon metabolism could have therapeutic potential in preserving cognition.

Conversely, genes with negative feature importance are enriched for inflammatory cytokine signaling, necroptotic cell death, and astrocyte activation programs. Key contributors such as *RIPK1*(55), which drives programmed necrotic cell death, and *IL6ST*(56) and *BIN1*(57), which regulate astrocyte differentiation and immune activation, reflect pathways consistently implicated in AD neuroinflammation and neuronal vulnerability. The prominence of these pathways underscores the central role of immune dysregulation and glial reactivity in accelerating cognitive decline. These results reinforce the notion that targeting neuroinflammatory circuits or necroptotic pathways may mitigate AD progression, particularly in individuals showing early molecular signatures of vulnerability.

At the metabolomic level, positive-direction metabolites overlap with obesity- and AD-associated metabolic signatures, including hexanoylcarnitine (C6)(58), benzoate(59), 3-phenylpropionate(60), imidazolelactate(61), and 3-methylhistidine(62). These metabolites collectively reflect a metabolic state characterized by efficient substrate utilization and adaptive energy metabolism, aligning with the cognitive resilience observed in WRAP participants. The convergence of these metabolites with protective profiles suggests that systemic metabolic efficiency may be a modifiable axis for promoting cognitive health, and that dietary or pharmacologic strategies enhancing ketone utilization or gut-derived metabolite balance could support resilience.

In contrast, metabolites with negative feature importance correspond to signatures of metabolic dysfunction, including disorders such as glutaric aciduria II, very long-chain acyl-CoA dehydrogenase deficiency, and type 2 diabetes. Species such as sphingomyelins(63,64), 1-stearoyl-2-arachidonoyl-GPC (18:0/20:4), 1-linoleoyl-2-arachidonoyl-GPC (18:2/20:4), beta-cyanoalanine, and cinnamyl alcohol point toward disrupted lipid handling, mitochondrial stress, and impaired energy metabolism. These metabolic perturbations mirror the vulnerability patterns identified in AD cohorts and emphasize the tight link between systemic metabolic health and neurodegenerative risk. Such findings support the idea that interventions targeting mitochondrial function, fatty acid oxidation, or lipid homeostasis may hold promise for reducing AD susceptibility.

Overall, integrating transcriptomic and metabolomic feature importance illuminates two complementary axes of cognitive resilience and vulnerability: one centered on energy efficiency, epigenetic stability, and metabolic flexibility, and another driven by inflammation, glial activation, and metabolic dysregulation. These results highlight potential multi-level intervention points, from enhancing mitochondrial and metabolic health to modulating neuroinflammatory pathways, and underscore the value of multi-omic approaches in identifying mechanisms that underlie heterogeneity in AD progression.

Several limitations of this study should be noted. First, the gene expression levels in this study were imputed based on genotypes rather than directly measured. The accuracy of the imputation depends on how precise the pre-trained models could capture true gene expression patterns, which vary strongly across genes and tissues. Therefore, although this approach enabled a more comprehensive analysis when measured transcriptomics data are unavailable, we acknowledge that it may introduce errors. Future studies could further improve accuracy by integrating more diverse reference datasets and refining the imputation models. Second, the use of the Wisconsin ADRC cohort as an independent validation set for the trained models may not provide a robust assessment of predictive performance due to its relatively small sample size. Utilizing larger independent cohorts with characteristics similar to WRAP and Wisconsin ADRC for validation would help improve the robustness of the models’ predictive performance. To further deepen our understanding of AD, integrating a wider array of biomarkers, such as those from proteomics, would provide a more holistic view of its pathology. Furthermore, while this study focuses on specific biomarkers and their pairwise interactions, AD is a complex and multifactorial disease. Therefore, other factors, such as environmental influences and epigenetic changes, which were not captured in this analysis, could also play critical roles in disease progression. In the future study, investigating the complex interplay between genetic, environmental, and epigenetic factors will offer valuable insights into the multifactorial nature of AD progression. Lastly, although this study primarily focused on individual feature importance, interactions between biomarkers may also play an important role in multi-omics regulation. Future work leveraging advanced multi-view integrative frameworks such as Cooperative multi-view integration with a scalable and interpretable model explainer (COSIME)(65) could enable scalable interpretation of across-omic feature interactions and provide deeper biological insight.

## Acknowledgement

The authors especially thank the WRAP and Wisconsin ADRC participants and staff for their contributions to the studies. Without their efforts, this research would not be possible.

## Author contributions

**Jerome J. Choi:** Conceptualization; Data curation; Formal analysis; Investigation; Methodology; Validation; Visualization; Writing – original draft; Writing – review & editing.

**Corinne D. Engelman:** Funding acquisition; Investigation; Writing – review & editing.

**Tianyuan Lu:** Conceptualization; Funding acquisition; Investigation; Writing – review & editing.

## Ethical considerations

Not applicable.

## Consent to participate

Not applicable.

## Consent for publication

Not applicable.

## Declaration of conflicting interest

The authors declare no competing interests.

## Funding

This study was supported by the National Institutes of Health (NIH) grants R01AG027161 (Wisconsin Registry for Alzheimer Prevention: Biomarkers of Preclinical AD), P30 AG062715 (the Wisconsin Alzheimer’s Disease Research Center), and RF1AG054047 (Genomic and Metabolomic Data Integration in a Longitudinal Cohort at Risk for Alzheimer’s Disease. Computational resources were supported by core grants from the Center for Demography and Ecology (P2CHD047873) and the Center for Demography of Health and Aging (P30AG017266). TL has been supported by start-up funding from the Department of Population Health Sciences, School of Medicine and Public Health, and Office of the Vice Chancellor for Research at the University of Wisconsin-Madison.

## Data availability statement

The data supporting the findings of this study are available on request from the corresponding author. The data are not publicly available due to privacy or ethical restrictions.

## Supplementary Information

**SUPPLEMENTARY FIGURE 1.**
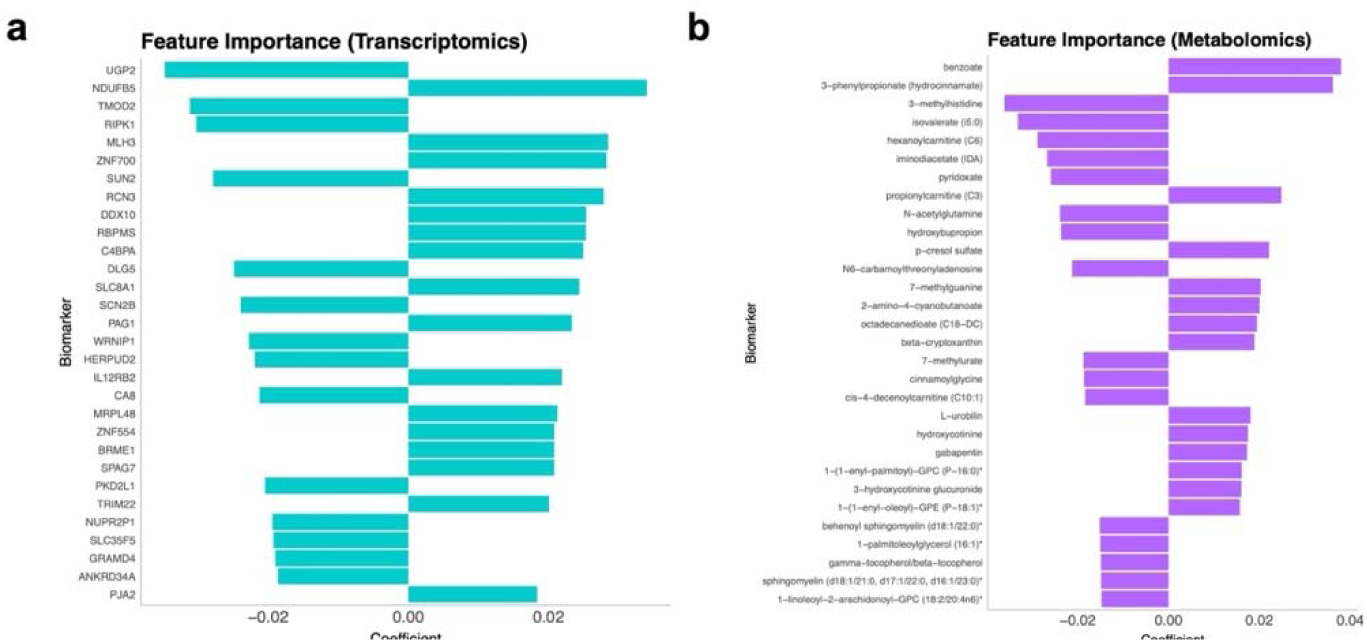
Biomarker importance values. **a** Gene importance values. **b** Metabolite importance values. Feature importance values correspond to model coefficients, with sign indicating the direction of association with PACC3. For each modality, the top 30 features ranked by the absolute value of their coefficients are shown.

## Notes

### Competing Interest Statement

The authors have declared no competing interest.

### Author Declarations

All study procedures were approved by the University of Wisconsin School of Medicine and Public Health Institutional Review Board and are in accordance with the Declaration of Helsinki.

